# A BIBLIOMETRIC ANALYSIS OF RHEUMATOLOGY AND COVID-19 RESEARCHES

**DOI:** 10.1101/2021.03.31.21254695

**Authors:** Ozge Pasin, Tugce Pasin

**Affiliations:** PhD, Department of Biostatistics, Faculty of Medicine, Istanbul University, Istanbul, Turkey; MD, Department of Physical Medicine and Rehabilitation, Istanbul Goztepe Training and Research Hospital, Istanbul, Turkey

**Author notes:** **Corresponding Author:** Ozge Pasin, PhD, Department of Biostatistics, Istanbul Faculty of Medicine, Istanbul University, Capa-Fatih Istanbul, TURKEY, E-mail address, /33412.

**Keywords:** COVID-19, Epidemiology, Health Services Research

## Abstract

**Objectives:** COVID-19 has had a substantial impact on rheumatology. This study provides a general overview of studies on rheumatology and COVID-19.

**Methods:** Data were taken from the Web of Science (WoS) website. Analysis and network visualization mapping processes were carried out using VOSviewer. A total of 234 publications were analyzed, and the correlations between citation numbers and reference counts, usage counts and page numbers were analyzed with Spearman correlation coefficients.

**Results:** The average number of citations per item was 6.03. The studies were cited 1,411 times in total, and 1,121 times without self-citations. The countries with the highest number of publications on rheumatology and COVID-19 were the USA and England; the countries with the highest number of citations were Italy and the USA, and Jinoos Yazdany was the most cited author. The Annals of the Rheumatic Diseases was the most cited journal, whereas the highest number of articles on rheumatology and COVID-19 were published in Arthritis and Rheumatology.

**Conclusions:** Bibliometric analysis of rheumatology and COVID-19 can be useful to future studies because it provides a general perspective on the studies. This study provides an insight into the development of publications on rheumatology during the COVID-19 pandemic.

## INTRODUCTION

SARS-CoV-2 (severe acute respiratory coronavirus 2 syndrome), a new member of the coronavirus family, was first detected in Wuhan, China in December 2019. The virus causes fever, cough, fatigue, loss of taste and smell, dyspnea, myalgia, vomiting, diarrhea and progressive diseases. Severe forms can cause acute respiratory distress syndrome (ARDS) and death, with interstitial lung involvement accompanied by alveolar damage [1-3].

Patients with rheumatism are considered to be a COVID-19 risk group. When patients with rheumatological diseases are diagnosed with a COVID-19 infection, they should immediately contact their rheumatologist. Rheumatologic diseases are heterogeneous. Corticosteroids, synthetic and biological disease-modifying antirheumatic drugs (DMARDs) increase the risk of disease aggravation because they cause immunosuppression [4]. It is essential to understand what is driving the increased risk of COVID-19-related deaths in rheumatologic patients during the pandemic. Hydroxychloroquine, used for the treatment of rheumatological diseases, was used in the prevention and treatment of COVID-19, but subsequent clinical trials have not found any benefit [5]. Cytokine inhibitor drugs, such as Interleukin-6 (IL-6) inhibitors, were investigated to determine their effectiveness in the prevention and treatment of COVID-19 infections and complications, including cytokine-storm [6].

Rheumatism patients are at risk of developing infections due to the disease itself, the drugs they use, and their more serious prognosis. The course of COVID-19 in individuals with rheumatological diseases could not be clearly determined because COVID-19 is a new and recently identified virus, and rheumatic diseases are less common than other diseases.

This study uses bibliometric methods to analyze publications on rheumatology and COVID-19.

## MATERIALS AND METHODS

The Web of Science (WoS) Core Collections was used to find publications for analysis. The documents were saved in ANSI format, and the bibliometric analysis and all other processes were carried out with VOSviewer (version 1.6.16). The dataset is open source, so there is no need for approval by an ethical committee. For the analysis, we used the following keywords: “COVID-19” and “Rheumatology”; “Coronavirus” and “Rheumatology”; “2019-nCoV” and “Rheumatology”; “SARS-CoV-2” and “Rheumatology”; “COVID-19” and “Rheumatic Disease”; “Coronavirus” and “Rheumatic Disease”; “2019-nCoV” and “Rheumatic Disease”; “SARS-CoV-2” and “Rheumatic Disease”; “COVID-19” and “Rheumatism”; “Coronavirus” and “Rheumatism”; “2019-nCoV” and “Rheumatism”; and “SARS-CoV-2” and “Rheumatism.” The analyzed works (234 in total) were published in 2020 and 2021 and include articles and abstracts. The relationships between citation numbers and some related variables were analyzed with Spearman correlation coefficients, and p-values are reported and tested at a significance level of 0.05 using IBM SPSS Statistics 21.

## RESULTS

The results show that 46.58% (109) of the documents were articles and 14.95% were meeting (seminers etc) articles. Table 1 shows the document types that were analyzed. The studies were cited 1,411 times in total, and 1,121 times without self-citations. Most publications, 216 (92.308%), were written and published in English, followed by 13 (5.556%) in German, and the remaining five (2.137%) were in Spanish.

**Table 1.** Publication Types

| Publication Type | Count | % |
| --- | --- | --- |
| Article | 109 | 46.581 |
| Meeting Abstract | 35 | 14.957 |
| Editorial Material | 33 | 14.103 |
| Review | 32 | 13.675 |
| Letter | 21 | 8.974 |
| Early Access | 17 | 7.265 |
| News Item | 4 | 1.709 |

Table 2 shows the top ten countries ranked by number of articles and citations. The highest number of articles were from the USA, England, Italy, Germany, Canada, Spain, Australia, Turkey, France, and New Zealand. The most cited publications were from Italy, with 672 citations.

**Table 2.** The top ten list of the number of articles and citations by country

| Country | Documents | Country | Citations |
| --- | --- | --- | --- |
| USA | 75 | Italy | 672 |
| England | 40 | USA | 454 |
| Italy | 30 | England | 300 |
| Germany | 24 | Canada | 293 |
| Canada | 23 | Spain | 255 |
| Spain | 18 | Australia | 240 |
| Australia | 14 | Germany | 236 |
| Turkey | 12 | France | 207 |
| France | 11 | New Zealand | 205 |
| New Zealand | 9 | Portugal | 190 |

Table 3 shows the top ten authors ranked by number of documents and citations. For the purpose of analysis, the minimum number of documents per author was set at five. only ten out of all the authors analyzed contributed to at least five publications. Jinoos Yazdany’s articles on rheumatology and COVID-19 were the most cited.

**Table 3.** The top ten list of the authors, journals and organizations

|  | Author | Documents | Citations | Journals | Documents | Citations | Organization | Documents | Citations |
| --- | --- | --- | --- | --- | --- | --- | --- | --- | --- |
| 1 | Jinoos Yazdany | 10 | 113 | Annals of the Rheumatic Diseases | 14 | 245 | University of California San Francisco | 13 | 234 |
| 2 | Emily Sirotych | 10 | 83 | Rheumatology International | 18 | 42 | The University of Queensland | 12 | 231 |
| 3 | Pedro M. Machado | 7 | 67 | Clinical Rheumatology | 16 | 32 | Harvard Medical School | 12 | 225 |
| 4 | Rebecca Grainger | 7 | 83 | Lancet Rheumatology | 11 | 104 | University of Otago | 8 | 205 |
| 5 | Philip Robinson | 5 | 66 | Seminars in Arthritis and Rheumatism | 5 | 37 | Canadian Arthritis Patient Alliance | 6 | 201 |
| 6 | Jonathan Hausmann | 5 | 0 | Arthritis and Rheumatology | 42 | 83 | Mc Master University | 10 | 201 |
| 7 | Philip C. Robinson | 5 | 47 | Zeitschrift für Rheumatologie | 11 | 11 | Massachusetts General Hospital | 9 | 167 |
| 8 | Roberto Caporali | 6 | 40 | Rheumatology | 6 | 15 | University of Washington | 8 | 209 |
| 9 | Hamdi Wafa | 5 | 5 | International Journal of Rheumatic Diseases | 7 | 4 | Healthpartners | 5 | 193 |
| 10 | Amy S. Mudano | 5 | 63 |  |  |  | UCL - London's Global University | 12 | 84 |

Figure 1 shows the network visualization map of international cooperation among countries that published articles on rheumatology and COVID-19. The size of the circle relates to the number of articles, colors indicate clusters, and line thickness is relative to the strength of the relationship. The minimum number of citations per country was eleven, and twenty countries satisfied this condition.

**Figure 1.**
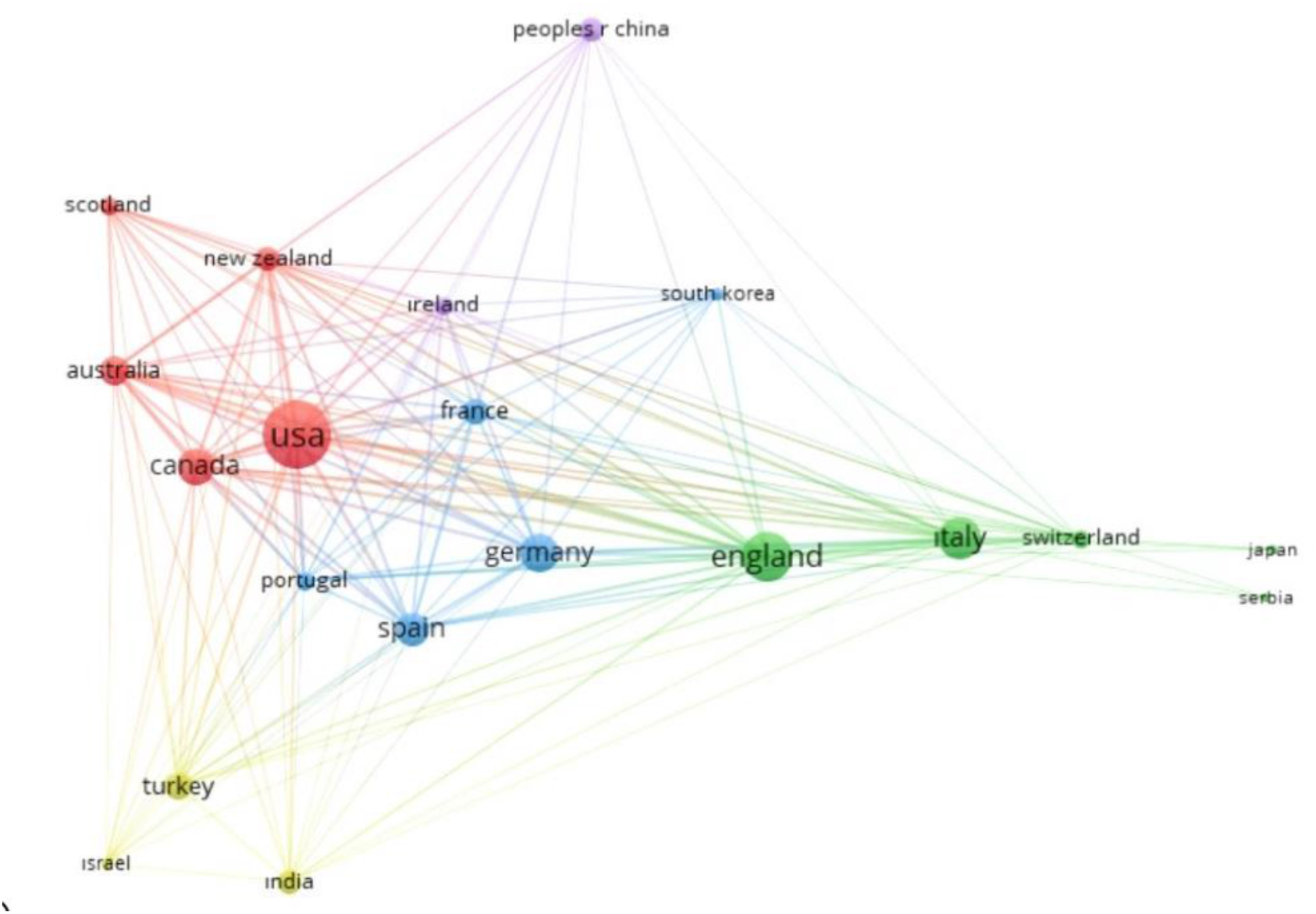
Network visualization map for international cooperation of world countries publishing publications on rheumatology and COVID-19

Table 3 lists journals ranked by most documents and citations. The minimum number of documents per journal was set at five, and nine journals met this criterium. The *Annals of the Rheumatic Diseases* was the most cited journal, whereas most articles on rheumatology and COVID-19 were published in *Arthritis and Rheumatology*.

Figure 2 shows a network visualization map for citations of current journals that publish articles on rheumatology and COVID-19. The size of the circle relates to the number of articles, colors indicate clusters, and line thickness is relative to the strength of the relationship. The minimum number of documents per organization was five, and 26 organizations satisfied this condition.

**Figure 2.**
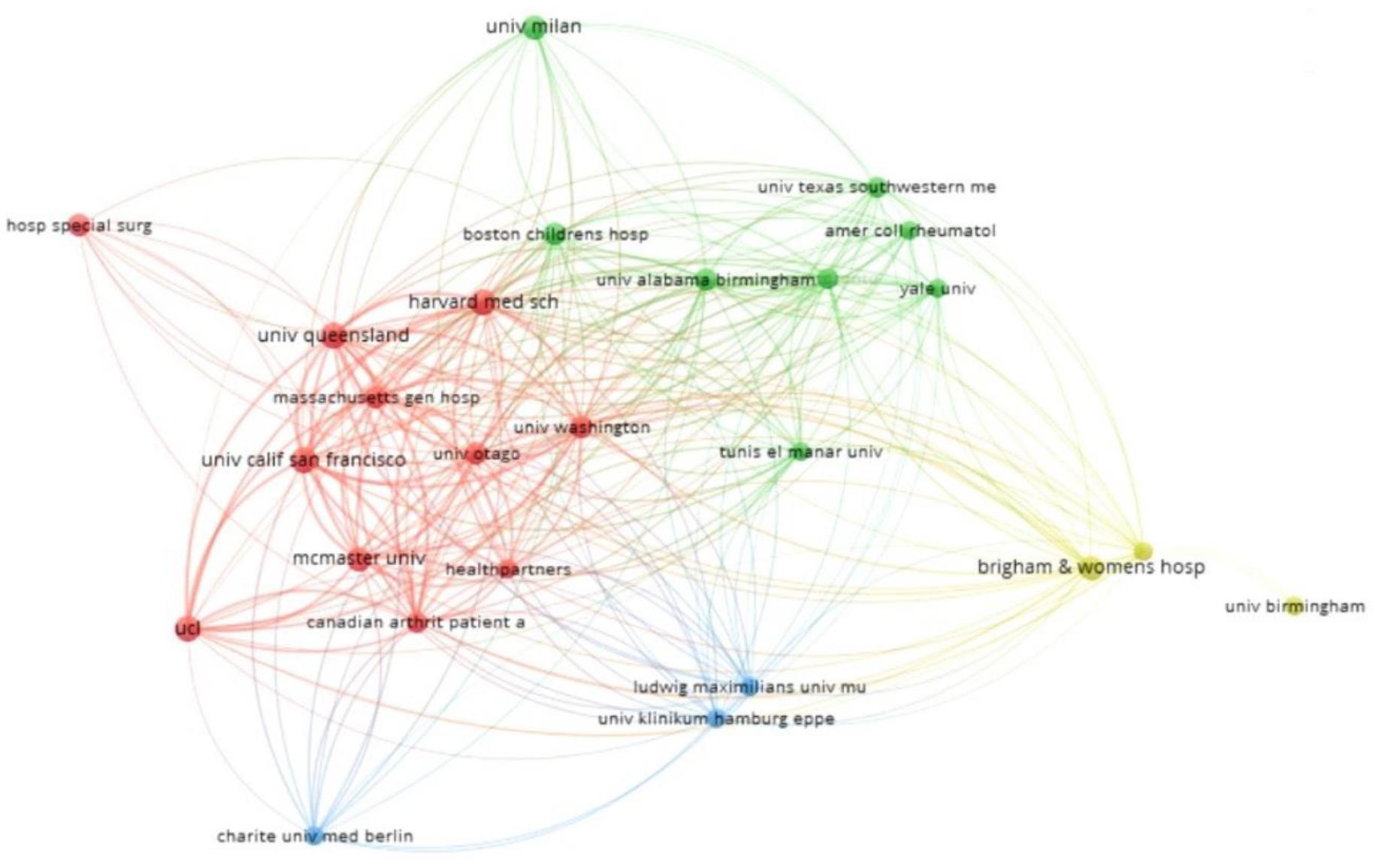
Network visualization map for citation analysis of active journals on rheumatology and COVID-19

Table 3 lists the organizations that submitted the most articles for publication. The minimum number of documents per organization was set at five. Taking into account the numbers of documents and citations, the ten most active institutions were the University of California San Francisco, The University of Queensland, Harvard Medical School, University of Otago, Canadian Arthritis Patient Alliance, McMaster University, Massachusetts General Hospital, University of Washington, Health Partners, and UCL.

The correlation between the number of citations and the 180-day usage count, the 2013 onwards usage count, and the number of pages is shown in Table 4. The usage count is a measure of the level of interest in a specific item on the WoS platform and reflects the number of times that an article has met a user’s information needs. The count is based on the number of clicks to expand the full-length article at the publisher’s website or on the number of downloads for use in a bibliographic management tool. The usage count is not a per-institution record—it is a record of all activity by all WoS users. The 180-day usage count is the number of times the full text of a record has been accessed or a record has been saved in the last six months. This count can increase or decrease as the timeline advances. The 2013 onwards usage count is the number of times the full text of a record has been accessed or a record has been saved since February 1, 2013. This count can increase or remain static over time [7].

**Table 4.** The relationship citations numbers between cited references count, 180 Day usage count, since 2013 usage count and number of pages

|  |  | Cited Reference Count | 180 Day Usage Count | Since 2013 Usage Count | Number of Pages |
| --- | --- | --- | --- | --- | --- |
| Times Cited, All Databases | Correlation Coefficient | 0.344 | 0.274 | 0.449 | 0.182 |
|  | p | <0.001 | <0.001 | <0.001 | 0.005 |
|  | N | 234 | 234 | 234 | 234 |

There were statistically significant relationships between the number of citations and the variables in the table (p < 0.001; p = 0.005). All the relationships were positive, but relationship strengths were low (r = 0.344; r = 0.274; r = 0.449; r = 0.182) (Table 4).

## DISCUSSION

Patients with rheumatism are at risk of developing infections due to the disease itself, the drugs they use, and their more serious prognosis during the pandemic. The course of COVID-19 in individuals with rheumatological diseases could not be clearly determined because COVID-19 is a new and recently identified virus, and rheumatic diseases are less common than other diseases.

The use of cortisone for the treatment of rheumatological diseases causes an increase in overall infection rates, and especially the rate of viral infections, depending on the dose, duration of treatment, and total dosage—but even at dosages considered safe, there is an increased risk of infection. Fredi and friends indicate that patients with rheumatic and musculoskeletal diseases do not appear to have a milder form of COVID-19 pneumonia than the controls [8]. Therefore, the aim of this study is to evaluate papers on rheumatology diseases and COVID-19. The bibliometric analysis summarized 234 publications on rheumatology and COVID-19, which provides an insight into publications and citations by organization, country and author. There is a limitation: the only source of publications was the WoS core.

To the best of our knowledge, this paper is the first bibliometric analysis on rheumatology and COVID-19, which could be useful for future studies.

## Data Availability

The data in the study can be obtained in WOS.

## Funding

No financial support was taken in this article.

## Competing interests

The authors declare that there is no other conflict of interest to disclose.

## Contributors

All coauthors contributed to the development of the study design. Pasin O contributed to the statistical analysis and interpretation. In thr reporting part of the study all authors contributed.

## Acknowledgements

None

## Ethical approval information

Not required.

## Data sharing statement

The data in the study was obtained from Web of Science core.

## Notes

### Competing Interest Statement

The authors have declared no competing interest.

### Funding Statement

No funding.

### Author Declarations

There no need to take a ethical approval. There was no attemption to humans or animals.

